## Supplementary material for "Prevalence of chronic kidney disease among young people living with HIV in Sub Saharan Africa: A systematic review and meta-analysis": Study Protocol

Systematic Review Protocol

| Title of the review | Prevalence of Chronic Kidney Disease Among Young People Living with HIV in Africa: A Systematic Review |
| --- | --- |
| **First reviewer** | Esther Nasuuna |
| **Other reviewers (with role/contribution in the review)** | Nicholus Nanyenya, Davis Kibirige, Helen Weiss |
| **Funding source** | INTE-COMM Project |
| **PROSPERO registration number** | CRD42022347588. |

| **Amendments to the protocol** | None were made |
| --- | --- |

1. **Background to review**

### Brief introduction to the subject of the review, including rationale for undertaking the review and overall aim

Chronic kidney disease (CKD) is on the increase globally and is fast becoming a public health concern. Young people living with HIV (YPLHIV) have a growing burden of CKD with a four-fold risk of developing CKD during their lifetime. This is due to poorly controlled HIV, HIV infection of an immature immune and renal system, high viremia and black race. Prevalence of CKD varies from 7 to 12% in different regions of the world and is mostly attributable to non-communicable diseases such as diabetes and hypertension in the general population. Previous studies have concentrated on the general paediatric population and adults in high income countries whose prevalence might be different from the YPLHIV. Whilst widespread access to ART has reduced the incidence of HIV associated kidney diseases, it is well known that poor HIV control leads to HIVAN. It is not well known if the incidence and prevalence has increased among the YPLHIV who are living longer with HIV and have poorly controlled HIV. Despite these predispositions, the prevalence of CKD in YPLHIV is not well known in African countries including Uganda. Therefore, we aim to describe the prevalence of chronic kidney disease among young people aged 10 to 24 years living with HIV in Africa.

**2.** **Specific objectives/questions the review will address**

1. What is the prevalence of chronic kidney disease among young people living with HIV in Africa?

| **3. a) Eligibility Criteria for including studies in the review**  If the PICOS format does not fit the research question of interest, please split up the question into separate concepts and put one under each heading | |
| --- | --- |
| i. **Population, or participants and conditions of interest** | Young people aged 10 to 24 years living with HIV |
| ii. **Interventions/Exposure/item of interest** | Diagnosis of chronic kidney disease by  1. GFR<60ml/min/1.73m^2^  2. Persistent proteinuria or Albuminuria for three or more months  3. Radiological manifestations for three or more months |
| iii. **Comparisons or control groups, if any** | N/A |
| iv. **Outcomes of interest** | Prevalence of CKD |
| v. **Setting** | Sub Saharan Africa |
| vi. **Study designs** | Cross-sectional, cohort, randomised controlled trials, non-randomised controlled trials. |
| vii. **Time Period** | From 2000 to present |

**3. b) Criteria for excluding studies not covered in inclusion criteria**

Any specific populations excluded, date range, language, whether abstracts or full text available, etc

- Systematic reviews, case reports, conference abstracts, RCT protocols

| **4. Search methods** | |
| --- | --- |
| Electronic databases & websites  Please list all databases that are to be searched and include the interface (eg NHS HDAS, EBSCO, OVID etc) and date ranges searched for each. | PubMed/Medline, EMBASE, Web of Science, African Journals Online (AJOL), African Index Medicus, Africa wide Information, |
| **Other methods used for identifying relevant research**  ie contacting experts and reference checking, citation tracking | Reference Checking of key papers, citation tracking, grey literature search (conference presentations and abstracts, unpublished manuscripts/preprints) |
| Journals hand searched  If any are to be hand searched, please list which journals and date searched from, including a rationale. | N/A |

| **5. Methods of review** | |
| --- | --- |
| How will search results be managed & documented?  i.e. which reference management software, how duplicates dealt with | 1. All the citations will be exported to endnote reference management software.  2. Duplicates will be deleted.  3. All remaining articles will then be exported to the Rayyan software (https://rayyan.qcri.org/welcome) to do the title and abstract search. This will inform the articles that get full text review. Rayyan will manage the inclusion/exclusion process at each stage of the review and facilitate the selection process by reviewers. |
| Selection process  Number of reviewers, how agreements to be reached and disagreements dealt with, etc. | The articles will be screened by the first reviewer (EMN) by title only, all duplicates and clearly inappropriate articles will be removed. The first (EMN), second (NN) and third reviewers will then each independently screen 50% of the remaining articles by abstract to ensure that each article is reviewed by two reviewers.  From this list of selected articles, the three reviewers will then undertake the same process (50% each) and independently select papers for inclusion based on their full text. Where arbitration is necessary on study inclusion or exclusion, the third reviewer who did not review that article will arbitrate. EMN & NN will independently extract the relevant information from the studies to include in the review, with the two independent data extraction sheets compared and finalised by EMN. |
| Quality assessment  Tools or checklists used with references or URLs, was this piloted? Is it to be carried out at same time as data extraction? | The Joanna Briggs Institute (JBI) checklist for prevalence studies will be used to assess the quality of the studies. GRADE will be used to summarise the quality of the evidence and the strength of recommendations |
| **How is data to be extracted?**  What information is to be collected on each included study? If databases or forms on Word or Excel are used, were these piloted and how is this recorded and by how many reviewers? | A data extraction form will be designed and used. A database will be created in Microsoft EXCEL where details of all the relevant studies will be extracted and stored.  The following data is to extracted into an Excel sheet.   - First author - Country - Region - Population - Study setting - Age distribution - Sex distribution - Sample size - Number of CKD patients - Disease definition/diagnosis - Prevalence - 95% confidence interval of the prevalence - Symptoms - Equation used - Follow up |
| **Outcomes to be extracted & hierarchy/priority of measures**  i.e. which measure is preferred and if that is not available which is next in order of preference? | Any reported outcome, these may include, but are not limited to:   - Prevalence of CKD - Proportion diagnosed with CKD - Frequency of CKD diagnosis |
| Narrative synthesis  Details of what methods, how synthesis will be done and by whom. Is the Narrative Synthesis Framework to be used? | The narrative synthesis framework will be used. |
| **Meta-analysis**  Details of what and how analysis and testing will be done. If no meta-analysis is to be conducted, please give reason. | Random effects meta-analysis will be conducted. The design of the studies will be assessed to determine if the prevalence of CKD can be meaningfully combined and reported as a meta-analysis. Heterogeneity will be determined using Cochrane’s Q and if it is above 50%, then the studies will not be combined and a narrative reported instead |
| Will the overall strength of evidence be assessed? If so, how?  i.e. GRADE? | N/A |

| **6. Presentation of results** | |
| --- | --- |
| Outputs from review  Papers and target journals, conference presentations, reports, etc | Manuscript for publication in Plos Global Public Health |

| **7. Timeline for review – when do you aim to complete each stage of the review** | |
| --- | --- |
| **Protocol** | June 2022 |
| **Literature searching** | July 2022 |
| **Screening search results** | July to August 2022 |
| **Quality appraisal** | August 2022 |
| **Data extraction** | August 2022 |
| **Synthesis** | September 2022 |
| **Writing up** | October 2022 |
