## Supplementary material for "Prevalence of chronic kidney disease among young people living with HIV in Sub Saharan Africa: A systematic review and meta-analysis": Final Search Terms

The key words for all the searches were chronic kidney disease, young people, HIV and Africa.

**Medline/PubMed Search Strategy**

**Chronic kidney disease**

Kidney Diseases/

Renal Insufficiency/

exp Renal Insufficiency, Chronic/

(chronic kidney OR chronic renal).ti,ab

(CKF OR CKD OR CRF OR CRD).mp.

Pre_dialy* or predialy*

(microalbuminuria OR macroalbuminuria OR albuminuria OR proteinuria).ti,ab.

glomerulo* OR glomerular.tw. OR GFR

nephriti* or nephro*.tw.

**HIV**

HIV Infections/

HIV/ or HIV2/ or HIV1/

HIV or hiv_1* or hiv_2* or hiv1 or hiv2 or hiv_I* or hiv_II* or hivI*

"human immunodeficiency virus" or "human immunedeficiency virus" or "human immune_deficiency virus" or "human immune_deficiency virus" or (human immun* and deficiency virus).ti,ab

"acquired immunodeficiency syndrome" or "acquired immunedeficiency syndrome" or AIDS OR "acquired immune_deficiency syndrome" or "acquired immune_deficiency syndrome"

**Young people terms**

1. "Young Adult*".mp. or Young Adult/

2.Young people OR Young person*

2. Adolescent OR Adolescen*

3. Youth*.mp.

4. Child*.mp.

5. (Paediatric* OR pediatric*).ti,ab.

**Africa**

| 1 | "Africa south of the Sahara"/ |
| --- | --- |
| 2 | ("Africa South of the Sahara" or sub-Saharan Africa or subSaharan Africa).ti,ab. |
| 3 | Central Africa.ti,ab. |
| 4 | Eastern Africa.ti,ab. |
| 5 | Southern Africa.ti,ab. |
| 6 | Western Africa.ti,ab. |
| 7 | Seychelles/ |
| 8 | Seychelles.ti,ab. |
| 9 | Benin/ |
| 10 | (Benin or Dahomey).ti,ab. |
| 11 | Burkina Faso/ |
| 12 | (Burkina Faso or Burkina Fasso or Upper Volta).ti,ab. |
| 13 | Burundi/ |
| 14 | (Burundi or Ruanda-Urundi).ti,ab. |
| 15 | Central African Republic/ |
| 16 | (Central African Republic or Ubangi-Shari).ti,ab. |
| 17 | Chad/ |
| 18 | Chad.ti,ab. |
| 19 | Democratic Republic Congo/ |
| 20 | (((Democratic Republic or DR) adj2 Congo) or Congo-Kinshasa or Belgian Congo or Zaire or Congo Free State).ti,ab. |
| 21 | Eritrea/ |
| 22 | Eritrea.ti,ab. |
| 23 | Ethiopia/ |
| 24 | (Ethiopia or Abyssinia).ti,ab. |
| 25 | Gambia/ |
| 26 | Gambia.ti,ab. |
| 27 | Guinea/ |
| 28 | (Guinea not (New Guinea or Guinea Pig* or Guinea Fowl or Guinea-Bissau or Portuguese Guinea or Equatorial Guinea)).ti,ab. |
| 29 | Guinea-Bissau/ |
| 30 | (Guinea-Bissau or Portuguese Guinea).ti,ab. |
| 31 | Liberia/ |
| 32 | Liberia.ti,ab. |
| 33 | Madagascar/ |
| 34 | (Madagascar or Malagasy Republic).ti,ab. |
| 35 | Malawi/ |
| 36 | (Malawi or Nyasaland).ti,ab. |
| 37 | Mali/ |
| 38 | Mali.ti,ab. |
| 39 | Mozambique/ |
| 40 | (Mozambique or Mocambique or Portuguese East Africa).ti,ab. |
| 41 | Niger/ |
| 42 | (Niger not (Aspergillus or Peptococcus or Schizothorax or Cruciferae or Gobius or Lasius or Agelastes or Melanosuchus or radish or Parastromateus or Orius or Apergillus or Parastromateus or Stomoxys)).ti,ab. |
| 43 | Rwanda/ |
| 44 | (Rwanda or Ruanda).ti,ab. |
| 45 | Sierra Leone/ |
| 46 | (Sierra Leone or Salone).ti,ab. |
| 47 | Somalia/ |
| 48 | (Somalia or Somaliland).ti,ab. |
| 49 | south sudan/ |
| 50 | South Sudan.ti,ab. |
| 51 | Tanzania/ |
| 52 | (Tanzania or Tanganyika or Zanzibar).ti,ab. |
| 53 | Togo/ |
| 54 | (Togo or Togolese Republic or Togoland).ti,ab. |
| 55 | Uganda/ |
| 56 | Uganda.ti,ab. |
| 57 | Angola/ |
| 58 | Angola.ti,ab. |
| 59 | Cameroon/ |
| 60 | (Cameroon or Kamerun or Cameroun).ti,ab. |
| 61 | Cape Verde/ |
| 62 | (Cape Verde or Cabo Verde).ti,ab. |
| 63 | Comoros/ |
| 64 | (Comoros or Glorioso Islands or Mayotte).ti,ab. |
| 65 | Congo/ |
| 66 | (Congo not ((Democratic Republic adj3 Congo) or congo red or crimean-congo)).ti,ab. |
| 67 | Cote d'Ivoire/ |
| 68 | (Cote d'Ivoire or Cote dIvoire or Ivory Coast).ti,ab. |
| 69 | eswatini/ |
| 70 | (eSwatini or Swaziland).ti,ab. |
| 71 | Ghana/ |
| 72 | (Ghana or Gold Coast).ti,ab. |
| 73 | Kenya/ |
| 74 | (Kenya or East Africa Protectorate).ti,ab. |
| 75 | Lesotho/ |
| 76 | (Lesotho or Basutoland).ti,ab. |
| 77 | Mauritania/ |
| 78 | Mauritania.ti,ab. |
| 79 | Nigeria/ |
| 80 | Nigeria.ti,ab. |
| 81 | "sao tome and principe"/ |
| 82 | (Sao Tome adj2 Principe).ti,ab. |
| 83 | Senegal/ |
| 84 | Senegal.ti,ab. |
| 85 | Sudan/ |
| 86 | (Sudan not South Sudan).ti,ab. |
| 87 | Zambia/ |
| 88 | (Zambia or Northern Rhodesia).ti,ab. |
| 89 | Zimbabwe/ |
| 90 | (Zimbabwe or Southern Rhodesia).ti,ab. |
| 91 | Botswana/ |
| 92 | (Botswana or Bechuanaland or Kalahari).ti,ab. |
| 93 | Equatorial Guinea/ |
| 94 | (Equatorial Guinea or Spanish Guinea).ti,ab. |
| 95 | Gabon/ |
| 96 | (Gabon or Gabonese Republic).ti,ab. |
| 97 | Mauritius/ |
| 98 | (Mauritius or Agalega Islands).ti,ab. |
| 99 | Namibia/ |
| 100 | (Namibia or German South West Africa).ti,ab. |
| 101 | South Africa/ |
| 102 | (South Africa or Cape Colony or British Bechuanaland or Boer Republics or Zululand or Transvaal or Natalia Republic or Orange Free State).ti,ab. |
| 103 | or/1-102 [ALL SUB-SAHARAN AFRICA COUNTRIES] |

**Pubmed search**

**Kidney terms**

((((((((Kidney Diseases/) OR (Renal Insufficiency/)) OR (exp Renal Insufficiency, Chronic/)) OR ((chronic kidney OR chronic renal).ti,ab)) OR ((CKF OR CKD OR CRF OR CRD).mp.)) OR (Pre_dialy* or predialy*)) OR ((microalbuminuria OR macroalbuminuria OR albuminuria OR proteinuria).ti,ab.)) OR (glomerulo* OR glomerular OR GFR)) OR (nephriti* or nephro*)

**HIV terms**

((("human immunodeficiency virus" or "human immunedeficiency virus" or "human immune_deficiency virus" or "human immune_deficiency virus" or (human immun* deficiency virus).ti,ab "acquired immunodeficiency syndrome" or "acquired immunedeficiency syndrome" or AIDS OR "acquired immune_deficiency syndrome" or "acquired immune_deficiency syndrome") OR (HIV or hiv_1* or hiv_2* or hiv1 or hiv2 or hiv_I* or hiv_II* or hivI*)) OR (HIV/ or HIV2/ or HIV1/)) OR (HIV Infections/)

Young people

((((("Young Adult*".mp. or Young Adult/) OR (Young people OR Young person*)) OR (Adolescent OR Adolescen*)) OR (Youth*)) OR (Child*)) OR ((Paediatric* OR pediatric*).ti,ab.)

Africa

**exp "Africa South of the Sahara"/ OR ("Africa South of the Sahara" or sub-Saharan Africa or subSaharan Africa).ti,ab. OR (Central Africa).ti,ab. OR (Eastern Africa).ti,ab. OR (Southern Africa).ti,ab. OR (Western Africa).ti,ab. OR Angola/ OR Angola OR Cameroon/ OR (Cameroon or Kamerun or Cameroun).ti,ab. OR Cape Verde/ OR (Cape Verde or Cabo Verde).ti,ab. OR Comoros/ OR (Comoros or Glorioso Islands or Mayotte).ti,ab. OR Congo/ OR (Congo not ((Democratic Republic adj3 Congo) or congo red or crimean-congo)).ti,ab. OR Cote d'Ivoire/ OR (Cote d'Ivoire or Cote dIvoire or Ivory Coast).ti,ab. OR Eswatini/ (eSwatini or Swaziland).ti,ab. OR Ghana/ OR (Ghana or Gold Coast).ti,ab. OR Kenya/ OR (Kenya or East Africa Protectorate).ti,ab. OR Lesotho/ OR (Lesotho or Basutoland).ti,ab. OR Mauritania/ OR Mauritania OR Nigeria/ OR Nigeria OR (Sao Tome adj2 Principe).ti,ab. OR Senegal/ OR Senegal OR Sudan/ OR (Sudan not South Sudan).ti,ab. OR Zambia/ OR (Zambia or Northern Rhodesia).ti,ab. OR Zimbabwe/ OR (Zimbabwe or Southern Rhodesia).ti,ab. OR Botswana/ OR (Botswana or Bechuanaland or Kalahari).ti,ab. OR Equatorial Guinea/ OR (Equatorial Guinea or Spanish Guinea).ti,ab. OR Gabon/ OR (Gabon or Gabonese Republic).ti,ab. OR Mauritius/ OR (Mauritius or Agalega Islands).ti,ab. OR Namibia/ OR (Namibia or German South West Africa).ti,ab. OR South Africa/ OR (South Africa or Cape Colony or British Bechuanaland or Boer Republics or Zululand or Transvaal or Natalia Republic or Orange Free State).ti,ab. OR Benin/ OR (Benin or Dahomey).ti,ab. OR Burkina Faso/ OR (Burkina Faso or Burkina Fasso or Upper Volta).ti,ab. OR Burundi/ OR (Burundi or Ruanda-Urundi).ti,ab. OR Central African Republic/ OR (Central African Republic or Ubangi-Shari).ti,ab. OR Chad/ OR Chad OR "Democratic Republic of the Congo"/ (((Democratic Republic or DR) adj2 Congo) or Congo-Kinshasa or Belgian Congo or Zaire or Congo Free State).ti,ab. OR Eritrea/ OR Eritrea OR Ethiopia/ OR (Ethiopia or Abyssinia).ti,ab. OR Gambia/ OR Gambia OR Guinea/ OR (Guinea NOT (New Guinea or Guinea Pig or Guinea Fowl or Guinea-Bissau or Portuguese Guinea or Equatorial Guinea)).ti,ab. OR Guinea-Bissau/ OR (Guinea-Bissau or Portuguese Guinea).ti,ab. OR Liberia/ OR Liberia OR Madagascar/ OR (Madagascar or Malagasy Republic).ti,ab. OR Malawi/ OR (Malawi or Nyasaland).ti,ab. OR Mali/ OR Mali OR Mozambique/ OR (Mozambique or Mocambique or Portuguese East Africa).ti,ab. OR Niger/ OR (Niger not (Aspergillus or Peptococcus or Schizothorax or Cruciferae or Gobius or Lasius or Agelastes or Melanosuchus or radish or Parastromateus or Orius or Apergillus or Parastromateus or Stomoxys)).ti,ab. OR Rwanda/ OR (Rwanda or Ruanda).ti,ab. OR Sierra Leone/ OR (Sierra Leone or Salone).ti,ab. OR Somalia/ OR (Somalia or Somaliland).ti,ab. OR South Sudan/ OR ( South Sudan).ti,ab. OR Tanzania/ OR (Tanzania or Tanganyika or Zanzibar).ti,ab. OR Togo/ OR (Togo or Togolese Republic or Togoland).ti,ab. OR Uganda/ OR Seychelles/**

### EMBASE Search Strategy

**Prevalence of Chronic kidney disease among young people living with HIV in Africa**

**Chronic kidney disease**

Kidney Diseases/

Renal Insufficiency/

exp Renal Insufficiency, Chronic/

chronic kidney or chronic renal

CKF or CKD or CRF or CRD

Pre_dialy$ or predialy$

microalbuminuria or macroalbuminuria or micro_albuminuria or macro_albuminuria or albuminuria or proteinuria

glomerulo$ OR glomerular OR GFR

nephriti$ or nephrotic

**HIV**

exp HIV Infections/ or "HIV infect*"

HIV/ or HIV2/ or HIV1/

HIV or hiv_1$ or hiv_2$ or hiv1 or hiv2 or hiv_I$ or hiv_II$ or hivI$

"human immunodeficiency virus" or "human immunedeficiency virus" or "human immune_deficiency virus" or "human immune_deficiency virus" or (human immun* and deficiency virus)

"acquired immunodeficiency syndrome" or "acquired immunedeficiency syndrome" or AIDS OR "acquired immuno‐deficiency syndrome" or "acquired immune‐deficiency syndrome"

**Young people terms**

Young people OR Young adult$ OR young person$

Adolescent OR Adolescen$

Youth$.mp. or juvenile/

child$/

Paediatric$ OR pediatric$

**Africa**

or/26-127 [ALL SUB-SAHARAN AFRICA COUNTRIES]

### African Wide Information

**Chronic kidney disease terms**

Kidney Diseases/

Renal Insufficiency/

chronic kidney or chronic renal

CKF or CKD or CRF or CRD

Pre_dialy* or predialy*

microalbuminuria or macroalbuminuria or albuminuria or proteinuria

glomerulo* OR glomerular OR GFR

nephriti* or nephro*

**Young people terms**

Young people* OR Young adult* OR young person*

Adolescent OR Adolescen*

Youth*

child*

Paediatric* OR pediatric*

**HIV terms**

"HIV infect*"

HIV/ or HIV2/ or HIV1/

HIV or hiv_1 or hiv_2 or hiv1 or hiv2 or hiv_I$ or hiv_II or hivI

"human immunodeficiency virus" or "human immunedeficiency virus" or "human immune_deficiency virus" or "human immune_deficiency virus" or (human immun* and deficiency virus)

"acquired immunodeficiency syndrome" or "acquired immunedeficiency syndrome" or AIDS OR "acquired immuno‐deficiency syndrome" or "acquired immune‐deficiency syndrome"

**Africa terms**

sub saharan africa or sub-saharan africa or sub sahara or sub-sahara

Africa

### African Journals online

site:www.ajol.info "chronic kidney disease" or proteinuria AND HIV
